# Graph theory analysis of induced neural plasticity post-Acceptance and Commitment Therapy for chronic pain

**DOI:** 10.1101/2020.10.19.20212605

**Authors:** Sarah K. Meier, Kimberly L. Ray, Noah C. Waller, Barry C. Gendron, Semra A. Aytur, Donald A. Robin

## Abstract

Chronic musculoskeletal pain affects the lives of over 50 million individuals in the United States, at a cost of more than $550 billion each year. Chronic pain leads to functional brain changes within those suffering from the condition. Not only does the primary pain network transform as the condition changes from acute to persistent pain, a state of hyper-connectivity also exists between the default mode, frontoparietal, and salience networks. Graph theory analysis has recently been used to investigate treatment-driven brain network changes. For example, current research suggests that Acceptance and Commitment Therapy (ACT) may reduce the chronic pain associated hyper-connectivity between the default mode, frontoparietal, and salience networks, as well as within the salience network. This study extended previous work by examining the associations between the three networks above and a meta-analytically derived pain network. Results indicate decreased connectivity within the pain network (including left putamen, right insula, left insula, and right thalamus) in addition to triple network connectivity changes after the four-week Acceptance and Commitment therapy intervention.

## 1. Introduction

Chronic musculoskeletal pain (CMP) affects the lives of over 50 million individuals in the United States at a rate of 11-40%, with contributions to medical costs and intervention programs being over $550 billion each year (Center for Disease Control and Prevention, 2018). CMP manifests after an eventual transition from acute pain, and central cognitive processes largely explain the persistent nature of pain in the instance of musculoskeletal damage (Mano et al., 2018). Co-morbid cognitive changes associated with chronic pain are often modulated by emotions (Reddan & Wager, 2018). These cognitive and emotional changes involve mood, overall psychological state, meaning-related thoughts, and the ability to learn new information. Thus, CMP commonly co-occurs with depression, anxiety, fatigue, difficulty remembering, and difficulty concentrating, resulting in substantial disabilities participating in work, social interactions, and self-care practices (Pitcher et al., 2019). These co-morbid effects are associated with anatomical and functional changes in specific brain circuitry and appear to alter the perception of and self-reflection about the pain itself (Bushnell et al., 2013).

Brain changes in CMP are widespread and involve the pain network and sensory, emotional, and cognitive control networks that process information (Morton et al., 2016). Areas commonly included in the pain network are the thalamus, the insular cortex, the primary and secondary somatosensory cortices, the anterior cingulate cortex, and the prefrontal cortex. It has been shown that areas of the pain network that deal with emotional and motivational modulations are active during pain (Morton et al., 2016). Not only is this pain network involved in CMP, the default mode (DMN), frontoparietal (FPN), and salience (SN) networks are as well.

Previous work has shown that chronic pain conditions impact the brain via networks noted above (Mitsi & Zachariou, 2016). The DMN is commonly hypothesized to be active when an individual is not participating in any given mental task. Increased posterior DMN connectivity proportional to the intensity of pain has previously been reported (Kuner & Flor, 2017). The FPN is typically featured in cognitive control aspects of neural processing. Data show that the FPN predicts how pain intensity will progress for those experiencing chronic pain, particularly within the first three months of the condition (Pfannmöller & Lotze, 2019). The SN is a prominent network in emotional control and social behavior especially in regards to detection of salient stimuli. The SN can also become active after painful stimuli, causing attention to shift away from the point of focus (Hemington et al., 2016). Hemington and colleagues also state that the SN abnormalities have been correlated with chronic pain, continuous pain stimuli, and pain-related symptomology.

Not only does each network independently demonstrate changes in functional connectivity in response to different types of chronic pain stimuli, it has been demonstrated that there is elevated functional connectivity among the DMN, FPN, and SN in individuals suffering from CMP (Cauda et al., 2014; Cottam et al., 2016; Doll et al., 2015; Hemington et al., 2016; Napadow et al., 2010; van Ettinger-Veenstra et al., 2019; Zhao et al., 2017). These three core networks, designated as the “triple network” in this paper, have become crucial in understanding higher levels of cognitive functioning and reasons it may become abnormal (Menon, 2011; Menon, 2018).

The current investigation was to extend Aytur and colleagues’ work (under review) on behavioral changes and neurological changes in connectivity within and between the DMN, FPN, and SN as a result of four weeks of Acceptance and Commitment Therapy (ACT) for chronic pain. Hayes and colleagues describe ACT as a “third generation approach” that is particularly sensitive to psychological phenomena and emphasizes contextual changes and directly targeted approaches to create a broad and flexible range of mental capabilities (Hayes et al., 2006). It is considered a “well-established” treatment for CMP by the American Psychological Association. This therapy focuses on 1) observing normally-occurring negative thoughts and feelings as they arise in one’s mind without trying to alter them in any way and 2) regularly behaving in accordance with personal values and life goals (Dahl & Lundgren, 2006). The acceptance technique of ACT has been shown to produce lower self-rated pain intensity, depression, pain-related anxiety, and pain avoidance while increasing physical, social, and occupational ability (Dahl & Lundgren, 2006). It is believed that through the mindfulness and acceptance techniques of ACT, psychological flexibility (an individual’s ability to continue participating in value-based behaviors during aversive experiences) increases while reducing responses to chronic pain sensations. These outcomes indicate a strong rationale for attempting the ACT practice with a CMP population.

Examining resting state network connectivity (in the DMN, FPN, SN, and pain network) of individuals with CMP who were exposed to an ACT intervention can provide insight into the neural substrates underlying the chronic pain experience. The Network Based Statistic (Zalesky et al., 2010) is the focus in the following evaluation of neurological changes pre- and post-ACT. The concept of graph theory is applicable here, as between-subject network comparison may reveal connectivity abnormalities. These abnormalities may be related to their CMP condition or the neurological changes induced post-ACT. Graph theory can be used to answer questions regarding the organization of functional connections across regions of the brain, the integration of information between the brain’s sub-networks, and the roles that certain brain regions may play in communication between themselves (Smith et al., 2013). Graph theory models the functional brain as a “network of networks”. Within these networks are “nodes”, representing specific regions of the brain, and “edges”, representing functional connections between two of those nodes. An advantage of this type of computational analysis is to provide strong visuals of the functional connectivity that exists within both healthy brains and dysfunctional brains in addition to providing quantitative measures of that connectivity data (Sporns, 2003).

Prior to this study, as well as Aytur and colleagues’ (2020) study, little was known about how the brain can change in response to specific interventions in certain disease states, such as CMP. The extent to which ACT interacts with the hypothesized neural systems underlying chronic pain was also unknown. The present study further investigated relationships between the default mode, frontoparietal, and salience networks. Specifically, this work extends the investigation of the triple network by also examining the role of a pain network (driven by a recent meta-analysis) in relation to CMP. While the triple network showed reduced connectivity previously, it is unknown how that affects potential change in the pain network. This work is important as subjective reports suggest decreased levels of perceived pain due to ACT, a treatment focusing on cognition and emotion rather than pain itself. As such, the primary hypothesis is that connectivity of the pain network should not change, however connectivity between the triple network and the pain network may change post-ACT since pain itself affects cognitive and emotional brain networks.

## 2. Methods

### 2.1. Participant Information

Nine female participants (age 47.59 years ± 16.54 years; eight right-handed) completed the entire four-week ACT protocol of Aytur and colleagues’ (2020) study after being recruited through community-based health care clinics and providing their informed consent. Participants were required to be at least 18 years of age, English-speaking, and to have had CMP for at least three months. Participants had to have an average pain interference score of at least four on question nine of the Brief Pain Inventory (Cleeland & Ryan, 1994). Ten participants began initial testing; however, one individual ended the baseline scan early due to a medical condition, was eliminated from the study at that time, and was prorated for their time in the study.

### 2.2. Neuropsychological/Behavioral Testing

Participants completed a series of neuropsychological assessments in their pre-treatment session in order to determine baseline measures of cognitive ability, quality of life, and pain level. Select domains from the NIH Cognition Battery, PROMIS (Patient-Reported Outcomes Measurement Information System; regarding pain interference, pain intensity, etc.), and Neuro-QoL™ (Quality of Life in Neurological Disorders; regarding sleep, depression, anxiety, etc.) were obtained from the NIH Toolbox and were administered via iPad. Any additional assessments were administered via paper copy. The full set of assessments (Supplemental Information, Table 1) was administered to each subject both pre- and post-ACT. Analysis methods and results for behavioral data are located in Supplemental Information.

### 2.3. rsfMRI Data Collection

Resting-state functional magnetic resonance imaging (rsfMRI) data were collected during two separate scan sessions using a Siemens Three Tesla (3T) Magnetom Prisma scanner at Boston University’s Cognitive Neuroimaging Center in Boston, MA, USA. The baseline MRI scan was collected before the treatment and a second MRI scan was collected within two weeks post-treatment. For each of these MRI visits, structural MP-RAGE images were collected (TR/TE= 2.53 s/1.32 ms, flip angle= 7°, field of view (FOV) = 256×320 mm, 0.8 mm^3^ resolution), followed by two eight-minute scans to obtain resting state functional images using a T2* weighted Echo Planar Imaging (EPI) sequence (2.5 mm^3^ resolution, 60 slices, TR/TE= 1.2 s/30 ms, 300 volumes, FOV= 205 mm, multi-slice interleaved ascending). All nine participants underwent this imaging protocol, and were simply instructed to lie still in the scanner with eyes open, fixating on the crosshair placed in their field of view. Prior to the scans, each participant completed an MRI safety screening to rule out any medical or other issues that may have jeopardized their safety or excluded them from the study.

### 2.4. Treatment Sessions

The entire implementation of ACT consisted of eight 90-minute twice weekly group sessions administered over four continuous weeks. The first four-week protocol included four of the female participants (ages 38-66) and the second four-week protocol included five of the female participants (ages 20-60). Each session was set up to consist of seven steps (see Supplemental Information, Figures 1 and 2). These two four-week ACT sessions were administered by two licensed recreational therapists who received training from an ACT specialist (J. Potter).

### 2.5. rsfMRI Data Preprocessing and Functional Connectivity Analysis

All imaging data collected from the fMRI scanner were preprocessed using standard approaches in Statistical Parametric Mapping software, version 12 (SPM12; Penny et al., 2006) implemented via MATLAB 9.3 (Higham & Higham, 2016; R2017b). First, all scan data were imported in the form of DICOM images and converted to Nifti files using the DICOM Import function in SPM12. Then, all functional data were realigned and co-registered to the standard Montreal Neuroimaging Institute (MNI) template in SPM12. Finally, the preprocessing steps of motion correction, slice-timing correction, normalization to remove any individual variability for between subject comparisons, and smoothing to increase signal to noise ratio were completed, also in SPM12. Various corrections for head movement were applied including smoothing using a 3-mm full-width at half maximum Gaussian kernel and a band-pass filter which preserved frequencies between 0.01 and 0.08 Hz.

Next, each participant’s brain data were parcellated into specific regions of interest (ROIs) for the DMN, FPN, and SN using a Multi-image Analysis GUI (Mango; Kochunov et al., 2002) selected from the Power atlas of 264 ROIs (Power et al., 2011). The mean time course (BOLD signal changes in time) within these seed regions was extracted from the residual images using Response Exploration for Neuroimaging Datasets (REX; Duff, 2008). Functional connectivity estimates across all selected ROIs were then calculated using the pairwise Pearson correlation of the region’s time course. Matrices were reduced to 101×101 ROIs for the combination of DMN, FPN, and SN (Figure 3A; see Supplemental Information, Table 2 for X, Y, Z coordinates of nodes in each of the four networks).

**Figure 3A.**
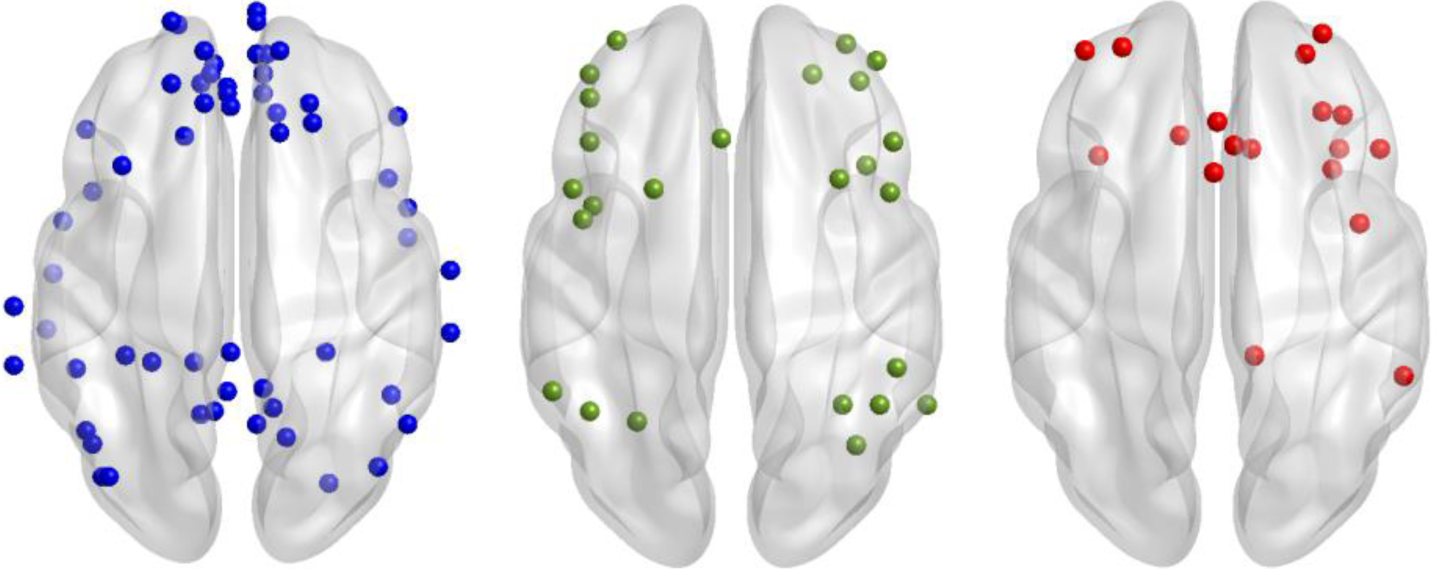
Original nodes used in each network (blue: DMN, green: FPN, red: SN).

### 2.6. Inclusion of Pain Network Nodes via Meta-Analysis

The pain matrix was created from a meta-analysis of regional activation in chronic pain patients (Waller et al., 2020). We chose to include functionally and meta-analytically derived nodes in order to be comparable to the functionally and meta-analytically derived Power atlas nodes. Waller and colleagues used the activation likelihood estimate (ALE; Eickhoff et al., 2012) to identify brain regions consistently activated during pain induction (for 419 subjects and 398 coordinate foci) and then applied meta-analytic connectivity modeling (MACM; Robinson et al., 2010) to yield co-activation patterns between those regions. This meta-analysis of chronic pain resulted in activation of seven brain areas (Figure 3B; right medial frontal gyrus, bilateral insula, right postcentral gyrus, left lentiform nucleus, right thalamus, and right claustrum). The seven pain network ROIs were included in network analyses as a fourth network of interest, resulting with a final connectivity matrix of 108×108 ROIs.

**Figure 3B.**
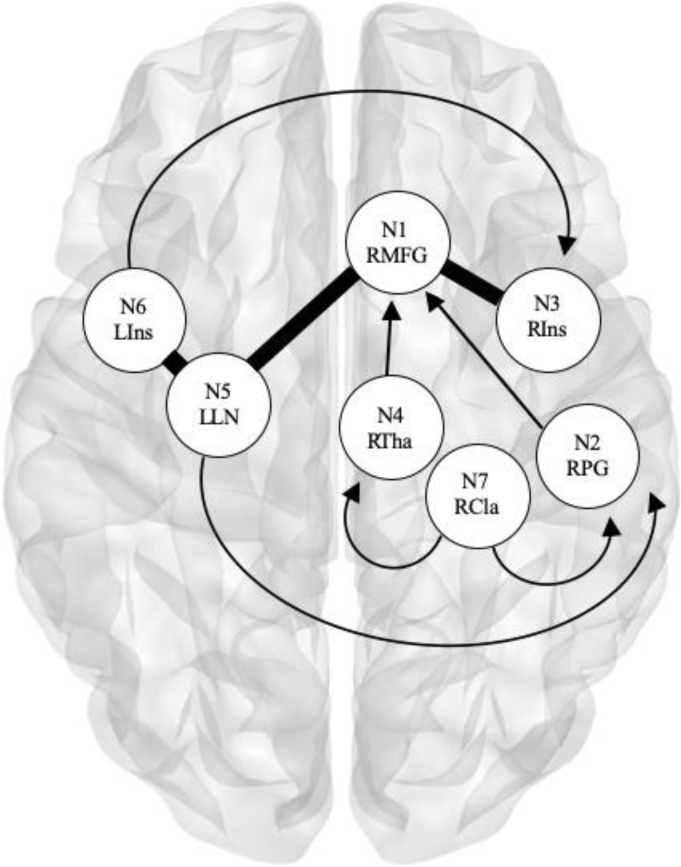
Pain nodes used, derived from chronic pain functional MACM results (Waller et al., 2020). ROI labels: N1 RMFG = right medial frontal gyrus, N2 RPG = right postcentral gyrus, N3 RIns = right insula, N4 RTha = right thalamus, N5 LLN = left lentiform nucleus, N6 LIns = left insula, N7 RCla = right claustrum. Arrows denote one-way connections projecting from the starting node. Thick non-arrows denote two-way connections between nodes.

### 2.7. rsfMRI Data Graph Theory Analysis

The Network Based Statistic (NBS; Zalesky et al., 2010) was used to evaluate the extent of the functional connectivity in the DMN, FPN, SN, and pain network pre- and post-ACT. Simply, this graph theory-based method utilizes a quantitative approach to identifying functional connectivity changes across selected regions by calculating parameters of a certain network and comparing those to a random network with the same number of nodes and edges. Results presented represent network-level functional connectivity differences for p > 0.05 at 10,000 permutations. While we predicted that we would observe decreases in connectivity as a result of ACT, NBS t-tests are one sided, therefore we tested for both increases and decreases in connectivity between pre- and post-ACT across a range of t-statistic thresholds.

In our initial report (Aytur et al., 2020) we tested for changes in brain networks underlying ACT-related outcomes in those with CMP using only triple network nodes without the inclusion of pain nodes (t > 2.1 for the 101 nodes of the DMN, FPN, and SN). The current work examines the ten nodes showing significant ACT treatment effects in Aytur and colleagues’ triple network connectivity analysis (detailed above) in addition to Waller and colleagues’ (2020) pain matrix of seven nodes found via ALE. A threshold of t > 2.5 was used for this t-test comparison of pre- versus post-ACT (17 nodes). We also tested for more widespread network effects in a follow up NBS analysis that included the original (101) triple network nodes in addition to Waller and colleagues’ seven pain nodes (108 total brain regions). The original 101 nodes were used to demonstrate connectivity found in the second analysis at the whole-network level. While significant effects were observed across a range of t-statistic thresholds, we present findings at a t-statistic threshold of t > 3.4 so that the number of nodes in the resultant network showing ACT effects were comparable to that of the original triple network analysis (Aytur et al., 2020).

## 3. Results

The present study investigated relationships between the default mode, frontoparietal, and salience networks by also examining the role of a pain network (driven by a recent meta-analysis) in relation to CMP. Through NBS, a range of t-statistic thresholds were used to determine functional connectivity differences across the aforementioned networks.

### 3.1. rsfMRI Connectivity Effects

Effects of ACT were observed by lowered levels of connectivity within the matrix of 101 triple network nodes during post-ACT compared to pre-ACT across 10 nodes and 10 edges corresponding to DMN, FPN, or SN when aggregated together (Figure 4A; t > 2.5 p = 0.05). This network is representative of the same finding in our previous report (Aytur et al., 2020). Subsequent analysis from the current investigation used these 10 nodes showing significant ACT treatment effects in our initial analysis in addition to the pain matrix of seven nodes (17 nodes total). From this, effects of ACT were observed by lowered levels of connectivity across seven nodes and six edges corresponding to DMN, SN, or the pain network when aggregated together (Figure 4B; t > 2.5 p = 0.004). Interestingly, no FPN nodes were involved in this significant network. The final, whole-network, analysis including the original 101 triple network nodes and the 7 pain nodes (108 nodes total) demonstrated effects of ACT by lowered levels of connectivity across 31 nodes and 34 edges corresponding to DMN, FPN, SN, or the pain network when aggregated together (Figure 4C; t > 3.4 p = 0.036). Nodes of the pain network that were involved in decreased functional connections include the left putamen, right insula, left insula, and right thalamus. No increases in connectivity were observed in any NBS test that would correspond to pre-ACT > post-ACT. Within network connectivity changes were represented by functional connections of color (a connection within SN nodes (red) will be represented by a red edge, e.g.).

**Figure 4.**
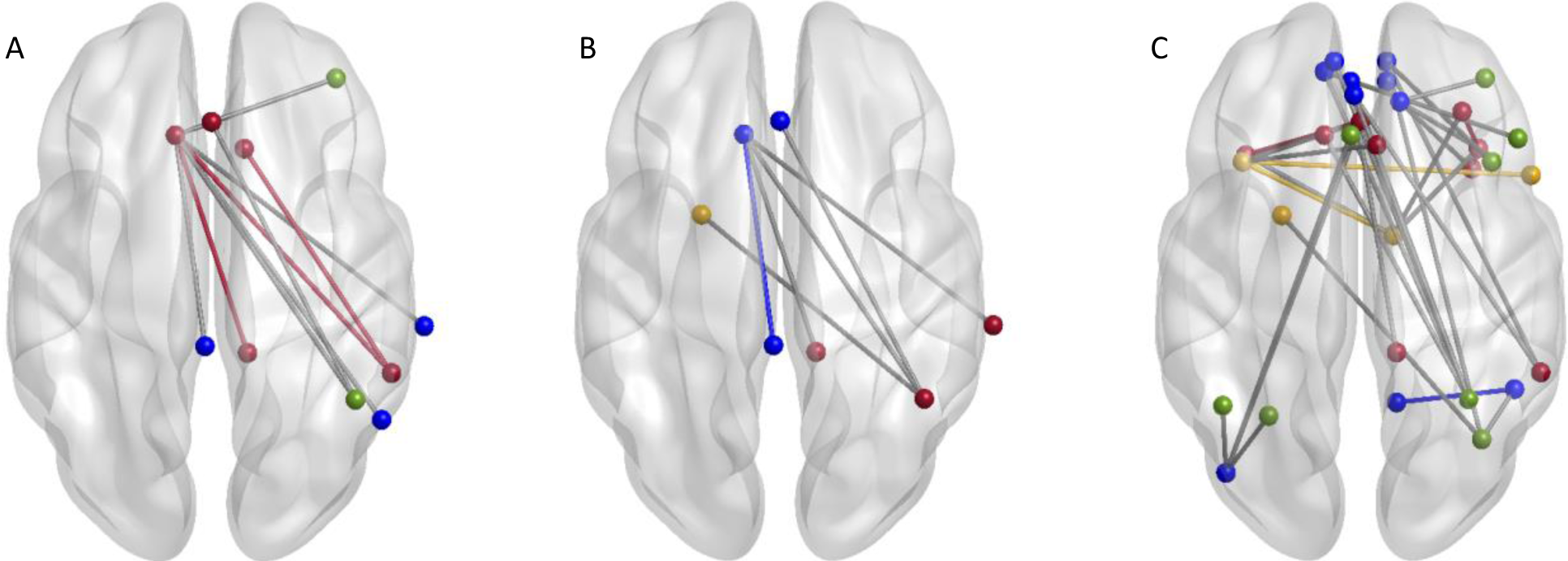
NBS identified resting state functional connectivity effects of ACT between DMN (blue), FPN (green), SN (red), and pain network (yellow). Showing decreased functional connectivity (post-ACT > pre-ACT) for A) 10 nodes connected by 10 edges (t >2.5; p = 0.05, 10,000 permutations), B) 7 nodes connected by 6 edges (t >2.5; p = 0.004, 10,000 permutations), C) 31 nodes connected by 34 edges (t >3.4; p = 0.036, 10,000 permutations). Figures were created using BrainNet (Xia, Wang, & He, 2013).

## 4. Discussion

The current study utilized functional connectivity analysis approaches to investigate the extent to which Acceptance and Commitment Therapy induces network-level changes in persons suffering from chronic pain. Previous studies have identified neurophysiological changes in the instance of chronic pain in the thalamus, insular cortex, anterior cingulate cortex, prefrontal cortex, as well as the primary somatosensory cortex and parietal cortex (Morton et al., 2016; Seminowicz et al., 2011). Such neural mechanisms have been linked to catastrophizing one’s pain (Zeidan et al., 2011). In other words, when the brain changes occur in these regions, negative patterns form leading to thoughts of helplessness, pessimism, and rumination. These sorts of thoughts are the root of the poorly perceived quality of life that individuals with chronic pain believe that they have. Supplemental information, along with current research (Aytur et al., 2020), describes strong, positive behavioral changes after ACT intervention for the participants.

Multiple previous studies also present the potential for alterations of the DMN (Napadow et al., 2010; Kornelsen et al., 2013; Kuner & Flor, 2017; Karafin et al., 2019; Wakaizumi et al., 2019; Zhang et al., 2019), FPN (Pfannmöller & Lotze, 2019; Kutch et al., 2017; Zheng et al., 2020; Androulakis et al., 2018), and SN (Hemington et al., 2016; Bishop et al., 2018; Seeley, 2019) in those experiencing chronic pain, exposing the necessity to further investigate neurological mechanisms underlying chronic pain. The hypotheses of Aytur and colleagues’ (2020) study were rooted in the main idea that hyper-connectivity between these three networks (DMN, FPN, and SN) exists in the brains of individuals with chronic pain (Hemington et al., 2016; Napadow et al., 2010; van Ettinger-Veenstra et al., 2019). Those results suggest that levels of heightened connectivity between the triple network returned to a more healthy, lower level of connectivity post-ACT. The current study utilized this information to guide the hypothesis that the cause may be the result of alterations to pain-related cognitive and emotional networks by examining relationships involving the pain network. Data demonstrating decreased functional connectivity involving pain network nodes support this idea.

### 4.1. Main Findings in rsfMRI Data

Network-based statistics were examined to determine functional connectivity changes within and between specific networks. Through examining the significant triple network (10 nodes from Aytur et al., 2020 initial investigation) in addition to the pain network of seven nodes (17 nodes total; see Figure 4B), involvement of the pain network was revealed. It is important to note that only connections in the DMN and SN demonstrate significantly decreased connectivity pre- to post-ACT for this analysis. This finding indicates the impact of DMN and SN dysfunction especially, and their involvement in the processing aspect of chronic pain conditions. It also designates DMN and SN as potential target networks for the Acceptance and Commitment Therapy. Subsequently, expanding the analysis to encompass the whole triple network in addition to the pain network (108 nodes total; see Figure 4C) revealed additional involvement of pain nodes as well as connections between each of the triple networks with decreases in functional connectivity from pre- to post-ACT. Four pain nodes displayed functional connectivity changes associated with ACT treatment. This unexpected finding suggests that connectivity between both the left putamen and right insula, as well as the left insula and right thalamus, of the pain network decreases pre- to post-ACT. Activation of these regions have been linked to thoughts of helplessness, pessimism, and rumination in the instance of CMP, as previously stated, so decreased functional connectivity in these regions may contribute to enhanced quality of life that is reported post-ACT (Zeidan et al., 2011; Aytur et al., 2020).

Changes in cognitive and emotional networks indicate the participants’ ability to deal with pain more efficiently after ACT intervention. Despite prior research suggesting that ACT can reduce pain symptomatology without altering the pain network, the current analyses do expose changes to the pain network as a result of ACT. This may be due to the use of ALE (Eickhoff et al., 2012) to define regions activated during chronic pain processing, whereas other (non-meta-analytically derived) nodal assignments may not be as well suited for chronic pain conditions specifically. These functional connectivity changes involving regions of the DMN, FPN, SN, and pain network (alongside previously reported positive behavioral outcomes, Aytur et al., 2020), are convincing underlying reasons for the way ACT effectively alleviates chronic pain symptomology.

## 5. Conclusions

This unique investigation examines the mechanism of action underlying an evidence-based treatment approach for chronic pain using novel graph theory analysis. Findings add to results of prior research suggesting that Acceptance and Commitment Therapy normalizes the hyper-connectivity that exists between the default mode, frontoparietal, and salience networks of individuals with chronic pain. This work also suggests that hyper-connectivity involving pain nodes is also alleviated. The current study contributes to the necessary knowledge of neurological mechanisms underlying chronic pain conditions as a means of optimizing the ACT treatment. Analyses of alternative treatments for CMP are important, as traditional pain medications do little to affect both the hyper-connectivity and dully perceived quality of life existing in those with chronic pain. Most common medications (painkillers, muscle relaxants, anti-depressants, e.g.) aim to reduce or manage the actual pain sensation, but can often lead to unwanted side effects and major life changes (Dahl & Lundgren, 2006). Non-pharmacologic treatments, such as ACT, need to continue to be considered as part of a multi-modal toolbox for pain management.

## Data Availability

Due to the nature of this research, participants of this study did not agree for their data to be shared publicly, so supporting data is not available.

