## Supplementary material for "Graph theory analysis of induced neural plasticity post-Acceptance and Commitment Therapy for chronic pain": SKM_NeuroimageClinical_finalsupp.pdf

Supplemental Information

**SI Table 1.** List of behavioral and neuropsychological assessments (administered pre- and post-treatment). *One of the assessments used in final correlational analysis (first pass) between behavioral data and rsfMRI functional connectivity data.

| Battery/Assessment Name | Domain | Approximate Duration |
|---|---|---|
| NIH Toolbox Cognition Battery (iPad): | | |
| List Sorting Working Memory (LSWM) | Working Memory | 7 minutes |
| *Pattern Comparison Processing Speed Test (PCPS) | Processing Speed | 3 minutes |
| • Measure of the amount of information that can be processed within a certain amount of time | | |
| Picture Sequence Memory Test (PSM) | Episodic Memory | 7 minutes |
| Flanker Inhibitory Control and Attention Test (FIC) | Attention, Executive Function | 3 minutes |
| Dimensional Change Card Sort Test (DCCS) | Executive Function | 4 minutes |
| Auditory Verbal Learning Test (AVL) | Episodic Memory | 3 minutes |
| Oral Symbol Digit Test (OSD) | Processing Speed | 3 minutes |
| Patient-Reported Outcomes Measurement Information System (PROMIS; iPad): | | |
| Pain Interference Survey | Pain | 1 minute |
| Pain Intensity Survey | Pain | 1 minute |
| Pain Behavior | Pain | 7 items |
| Severity of Substance Use | Substance Abuse | 7 items |
| Appeal of Substance Use | Substance Abuse | 7 items |
| Prescription Pain Medication Misuse | Substance Abuse | 7 items |
| Quality of Life in Neurological Disorders (Neuro QoL/NQ; iPad) - Short Form: | | |
| *Satisfaction with Social Roles and Activities (SSR) | Quality of Life | 25 minutes total: |
| • Measure of social health regarding satisfaction with involvement in one’s normal social roles, activities, or responsibilities | | |
| Stigma | Quality of Life | |
| Positive Affect and Well-Being (PA) | Quality of Life | |
| *Fatigue | Quality of Life | |
| • Measure of physical health regarding tiredness/exhaustion that decreases one’s functioning | | |
| Emotional and Behavioral Dyscontrol (EBD) | Quality of Life | |

| | |
|---|---|
| *Depression | Quality of Life |
| • Measure of mental health regarding feelings of hopelessness, negative mood, decrease in positive affect, negative self-views, etc. | |
| Anxiety | Quality of Life |
| Ability to Participate in Social Roles and Activities (APSR) | Quality of Life |
| Communication | Quality of Life |

| Additional Assessments (paper): | | |
|---|---|---|
| Current Opioid Misuse Measure (COMM) | Substance Abuse | 17 items |
| *Center for Epidemiologic Studies Depression Scale (CESD) | Depression | 20 items |
| • Measure of symptomology associated with depression | | |
| *PTSD Checklist (PCL-5) | PTSD | 20 items |
| • Measure of DSM-5 symptomology of post-traumatic stress disorder (PTSD) | | |
| Coping Strategies Questionnaire (CSQ) | Cognition | 45 items |
| Barratt Impulsiveness Scale (BIS-11) | Impulsivity | 30 items |
| *Brief Pain Inventory (BPI) | Pain | 9 items |
| • Measure of pain severity and its impact on functioning | | |
| *Acceptance & Action Questionnaire (AAQ-II) | Pain | 10 items |
| • Measure of ACT's behavioral effectiveness | | |
| Chronic Pain Acceptance Questionnaire (CPAQ) | Pain | 20 items |
| *Five Facet Mindfulness Questionnaire (FFMQ) | Mindfulness | 39 items |
| • Measure of five aspects of mindfulness: observing, describing, acting with awareness, non-judging, non-reacting | | |
| Dissociative Experiences Scale - II (DES-II) | Cognition | 28 items |

Approximate Total Time: 2 hours

| | | | | | | |
|---|---|---|---|---|---|---|
| Check in & set agenda | Review homework | Introduce the skill | Skill demonstration | In-session practice with feedback | Skill review | New homework |

**SI Figure 1.** The seven-step timeline of each individual 90-minute ACT session. “Homework” involved continuing practice of the skill learned in the prior session, as well as completing self-monitoring and self-reporting activities.

| | | | |
|---|---|---|---|
| Screening:<br><br>Eligibility, MRI screening | Pre-treatment scan and assessments:<br><br>Questionnaires for pain, cognitive measures, etc.<br><br>30-minute fMRI scan | ACT Treatment:<br><br>Two 90-minute sessions/week for four weeks | Post-treatment scan and assessments:<br><br>Questionnaires for pain, cognitive measures, etc.<br><br>30-minute fMRI scan |

**SI Figure 2.** A basic timeline of the entire study, starting with pre-treatment screening and ending with post-treatment assessments.

**SI Table 2.** List of all X, Y, Z coordinates for the nodes involved in each of the four networks. The ROI labels are directly from the Power atlas, except for those of the pain network (Power et al., 2011). Pain nodes were derived from Waller et al., 2020 meta analytic connectivity modelling.

| Default Mode Network | | | | Frontoparietal Network | | | |
|---|---|---|---|---|---|---|---|
| **ROI** | **X** | **Y** | **Z** | **ROI** | **X** | **Y** | **Z** |
| **74** | -41 | -75 | 26 | **174** | -44 | 2 | 46 |
| **75** | 6 | 67 | -4 | **175** | 48 | 25 | 27 |
| **76** | 8 | 48 | -15 | **176** | -47 | 11 | 23 |
| **77** | -13 | -40 | 1 | **177** | -53 | -49 | 43 |
| **78** | -18 | 63 | -9 | **178** | -23 | 11 | 64 |
| **79** | -46 | -61 | 21 | **179** | 58 | -53 | -14 |
| **80** | 43 | -72 | 28 | **180** | 24 | 45 | -15 |
| **81** | -44 | 12 | -34 | **181** | 34 | 54 | -13 |
| **82** | 46 | 16 | -30 | **186** | 47 | 10 | 33 |
| **83** | -68 | -23 | -16 | **187** | -41 | 6 | 33 |
| **86** | -44 | -65 | 35 | **188** | -42 | 38 | 21 |
| **87** | -39 | -75 | 44 | **189** | 38 | 43 | 15 |
| **88** | -7 | -55 | 27 | **190** | 49 | -42 | 45 |
| **89** | 6 | -59 | 35 | **191** | -28 | -58 | 48 |
| **90** | -11 | -56 | 16 | **192** | 44 | -53 | 47 |
| **91** | -3 | -49 | 13 | **193** | 32 | 14 | 56 |
| **92** | 8 | -48 | 31 | **194** | 37 | -65 | 40 |
| **93** | 15 | -63 | 26 | **195** | -42 | -55 | 45 |
| **94** | -2 | -37 | 44 | **196** | 40 | 18 | 40 |
| **95** | 11 | -54 | 17 | **197** | -34 | 55 | 4 |
| **96** | 52 | -59 | 36 | **198** | -42 | 45 | -2 |
| **97** | 23 | 33 | 48 | **199** | 33 | -53 | 44 |
| **98** | -10 | 39 | 52 | **200** | 43 | 49 | -2 |
| **99** | -16 | 29 | 53 | **201** | -42 | 25 | 30 |
| **100** | -35 | 20 | 51 | **202** | -3 | 26 | 44 |
| **101** | 22 | 39 | 39 | Salience Network | | | |
| **102** | 13 | 55 | 38 | **ROI** | **X** | **Y** | **Z** |
| **103** | -10 | 55 | 39 | **203** | 11 | -39 | 50 |
| **104** | -20 | 45 | 39 | **204** | 55 | -45 | 37 |
| **105** | 6 | 54 | 16 | **205** | 42 | 0 | 47 |
| **106** | 6 | 64 | 22 | **206** | 31 | 33 | 26 |
| **107** | -7 | 51 | -1 | **207** | 48 | 22 | 10 |
| **108** | 9 | 54 | 3 | **208** | -35 | 20 | 0 |

| | | | |
|---|---|---|---|
| **109** | -3 | 44 | -9 |
| **110** | 8 | 42 | -5 |
| **111** | -11 | 45 | 8 |
| **112** | -2 | 38 | 36 |
| **113** | -3 | 42 | 16 |
| **114** | -20 | 64 | 19 |
| **115** | -8 | 48 | 23 |
| **116** | 65 | -12 | -19 |
| **117** | -56 | -13 | -10 |
| **118** | -58 | -30 | -4 |
| **119** | 65 | -31 | -9 |
| **120** | -68 | -41 | -5 |
| **121** | 13 | 30 | 59 |
| **122** | 12 | 36 | 20 |
| **123** | 52 | -2 | -16 |
| **124** | -26 | -40 | -8 |
| **125** | 27 | -37 | -13 |
| **126** | -34 | -38 | -16 |
| **127** | 28 | -77 | -32 |
| **128** | 52 | 7 | -30 |
| **129** | -53 | 3 | -27 |
| **130** | 47 | -50 | 29 |
| **131** | -49 | -42 | 1 |
| **137** | -46 | 31 | -13 |
| **139** | 49 | 35 | -12 |

| | | | |
|---|---|---|---|
| **209** | 36 | 22 | 3 |
| **210** | 37 | 32 | -2 |
| **211** | 34 | 16 | -8 |
| **212** | -11 | 26 | 25 |
| **213** | -1 | 15 | 44 |
| **214** | -28 | 52 | 21 |
| **215** | 0 | 30 | 27 |
| **216** | 5 | 23 | 37 |
| **217** | 10 | 22 | 27 |
| **218** | 31 | 56 | 14 |
| **219** | 26 | 50 | 27 |
| **220** | -39 | 51 | 17 |
| Pain Network | | | |
| **ROI** | **X** | **Y** | **Z** |
| **1** | -36 | 18 | 2 |
| **2** | -24 | 2 | -2 |
| **3** | 40 | -14 | 6 |
| **4** | 52 | 14 | -2 |
| **5** | 4 | 16 | 44 |
| **6** | 58 | -18 | 18 |
| **7** | 10 | -4 | 2 |

**SI Table 3.** List of (three) significant edges of the salience network involving four nodes. The node assignment is listed out of the 264 Power atlas nodes. The brain regions corresponding to the nodes of each connection are listed also.

| **Power ROI** | **Corresponding Brain Regions:** |
|---|---|
| 203, 212 | Right medial cingulate cortex (MCC), Left anterior cingulate cortex (ACC) |
| 204, 212 | Right supramarginal gyrus (SupMG), Left anterior cingulate cortex (ACC) |
| 204, 217 | Right supramarginal gyrus (SupMG), Right anterior cingulate cortex (ACC) |

**SI Table 4.** List of the (34) significant edges (abbreviations) of triple network (t>3.4) found in Figure 6 with corresponding brain regions. The node assignment is listed out of the 264 Power atlas nodes. Items listed in **blue** are from Waller et al.'s (2020) pain ALE.

| ROI List | Abbreviation | Corresponding Brain Regions |
|---|---|---|
| 3, 35 | R MFG, R MFG | R medial frontal gyrus, R medial frontal gyrus |
| 36, 40 | L ACC, L SFG | L anterior cingulate, L superior frontal gyrus |
| 20, 55 | R Pre, R Ang | R precuneus, R angular gyrus |
| 36, 60 | L ACC, R IFG | L anterior cingulate, R inferior frontal gyrus |
| 47, 70 | R ACC, R MFG | R anterior cingulate, R medial frontal gyrus |
| 1, 72 | L MOcc, L SPL | L medial occipital lobe, L superior parietal lobule |
| 3, 75 | R MFG, R Ang | R medial frontal gyrus, R angular gyrus |
| 55, 75 | R Ang, R Ang | R angular gyrus, R angular gyrus |
| 1, 76 | L MOcc, L IPL | L medial occipital lobe, L inferior parietal lobule |
| 3, 77 | R MFG, R MFG | R medial frontal gyrus, R medial frontal gyrus |
| 36, 80 | L ACC, R IPL | L anterior cingulate, R inferior parietal lobule |
| 37, 80 | L SFG, R IPL | L superior frontal gyrus, R inferior parietal lobule |
| 38, 80 | L ACC, R IPL | L anterior cingulate, R inferior parietal lobule |
| 40, 80 | L SFG, R IPL | L superior frontal gyrus, R inferior parietal lobule |
| 1, 83 | L MOcc, L SFG | L medial occipital lobe, L superior frontal gyrus |
| 38, 84 | L ACC, R MCC | L anterior cingulate, R medial cingulate |
| 3, 85 | R MFG, R SMG | R medial frontal gyrus, R supramarginal gyrus |
| 37, 85 | L SFG, R SMG | L superior frontal gyrus, R supramarginal gyrus |
| 47, 90 | R ACC, R Ins | R anterior cingulate, R insula |
| 87, 90 | R MFG, R Ins | R medial frontal gyrus, R insula |
| 47, 92 | R ACC, R Ins | R anterior cingulate, R insula |
| 90, 92 | R Ins, R Ins | R insula, R insula |
| 80, 93 | R IPL, L ACC | R inferior parietal lobule, L anterior cingulate |
| 1, 96 | L MOcc, L ACC | L medial occipital lobe, L anterior cingulate |
| 80, 96 | R IPL, L ACC | R inferior parietal lobule, L anterior cingulate |
| 89, 96 | L Ins, L ACC | L insula, L anterior cingulate |
| 96, **102** | L ACC, **L Ins** | L anterior cingulate, **L insula** |
| 97, **102** | R MCC, **L Ins** | R medial cingulate, **L insula** |
| 75, **103** | R Ang, **L Put** | R angular gyrus, **L putamen** |
| **102, 105** | **L Put, R Ins** | **L putamen, R insula** |
| 87, **108** | R MFG, **R Thal** | R medial frontal gyrus, **R thalamus** |
| 89, **108** | L Ins, **R Thal** | L insula, **R thalamus** |
| 90, **108** | R Ins, **R Thal** | R insula, **R thalamus** |
| **102, 108** | **L Ins, R Thal** | **L insula, R thalamus** |

**SI Table 5.** List of (56) significant edges of the triple network (t>2.1). The node assignment is listed out of the 264 Power atlas nodes. The brain regions corresponding to the nodes of each connection are listed also, with the abbreviations used in this paper.

| ROI List | Corresponding Power Atlas Brain Regions |
|---|---|
| 96, 104 | Right angular gyrus (AG), Left superior frontal gyrus (SFG) |
| 76, 124 | Right medial frontal gyrus (orbital, MFG), Left parahippocampal gyrus (PHG) |
| 111, 124 | Left anterior cingulate cortex (ACC), Left parahippocampal gyrus (PHG) |
| 116, 124 | Right medial temporal gyrus (MTG), Left parahippocampal gyrus (PHG) |
| 123, 124 | Right medial temporal gyrus (MTG), Left parahippocampal gyrus (PHG) |
| 76, 126 | Right medial frontal gyrus (orbital, MFG), Left fusiform gyrus (FG) |
| 80, 126 | Right medial occipital gyrus (MOG), Left fusiform gyrus (FG) |
| 123, 126 | Right medial temporal gyrus (MTG), Left fusiform gyrus (FG) |
| 96, 127 | Right angular gyrus (AG), Right cerebellum (Cer) |
| 124, 174 | Left parahippocampal gyrus (PHG), Left precentral gyrus (PCG) |
| 124, 175 | Left parahippocampal gyrus (PHG), Right inferior frontal gyrus (triangular, IFG) |
| 124, 176 | Left parahippocampal gyrus (PHG), Left inferior frontal gyrus (opercular, IFG) |
| 126, 176 | Left fusiform gyrus (FG), Left inferior frontal gyrus (opercular, IFG) |
| 124, 178 | Left parahippocampal gyrus (PHG), Left superior frontal gyrus (SFG) |
| 126, 178 | Left fusiform gyrus (FG), Left superior frontal gyrus (SFG) |
| 124, 180 | Left parahippocampal gyrus (PHG), Right superior frontal gyrus (orbital, SFG) |
| 126, 180 | Left fusiform gyrus (FG), Right superior frontal gyrus (orbital, SFG) |
| 175, 180 | Right inferior frontal gyrus (triangular, IFG), Right superior frontal gyrus (orbital, SFG) |
| 139, 181 | Right inferior frontal gyrus (orbital, IFG), Right medial frontal gyrus (orbital, MFG) |
| 124, 188 | Left parahippocampal gyrus (PHG), Left medial frontal gyrus (MFG) |
| 126, 188 | Left fusiform gyrus (FG), Left medial frontal gyrus (MFG) |
| 180, 189 | Right superior frontal gyrus (orbital, SFG), Right medial frontal gyrus (MFG) |
| 124, 191 | Left parahippocampal gyrus (PHG), Left superior parietal gyrus (SPG) |
| 124, 192 | Left parahippocampal gyrus (PHG), Right inferior parietal lobule (IPL) |

| | |
|---|---|
| 126, 192 | Left fusiform gyrus (FG), Right inferior parietal lobule (IPL) |
| 188, 194 | Left medial frontal gyrus (MFG), Right angular gyrus (AG) |
| 124, 199 | Left parahippocampal gyrus (PHG), Right inferior parietal lobule (IPL) |
| 124, 201 | Left parahippocampal gyrus (PHG), Left inferior frontal gyrus (triangular, IFG) |
| 126, 201 | Left fusiform gyrus (FG), Left inferior frontal gyrus (triangular, IFG) |
| 180, 205 | Right superior frontal gyrus (orbital, SFG), Right precentral gyrus (PCG) |
| 124, 207 | Left parahippocampal gyrus (PHG), Right inferior frontal gyrus (triangular, IFG) |
| 180, 207 | Right superior frontal gyrus (orbital, SFG), Right inferior frontal gyrus (triangular, IFG) |
| 180, 208 | Right superior frontal gyrus (orbital, SFG), Left insula (Ins) |
| 124, 209 | Left parahippocampal gyrus (PHG), Right insula (Ins) |
| 205, 209 | Right precentral gyrus (PCG), Right insula (Ins) |
| 124, 210 | Left parahippocampal gyrus (PHG), Right inferior frontal gyrus (orbital, IFG) |
| 126, 210 | Left fusiform gyrus (FG), Right inferior frontal gyrus (orbital, IFG) |
| 94, 212 | Left medial cingulate cortex (MCC), Left anterior cingulate cortex (ACC) |
| 96, 212 | Right angular gyrus (AG), Left anterior cingulate cortex (ACC) |
| 119, 212 | Right medial temporal gyrus (MTG), Left anterior cingulate cortex (ACC) |
| 124, 212 | Left parahippocampal gyrus (PHG), Left anterior cingulate cortex (ACC) |
| 126, 212 | Left fusiform gyrus (FG), Left anterior cingulate cortex (ACC) |
| 189, 212 | Right medial frontal gyrus (MFG), Left anterior cingulate cortex (ACC) |
| 192, 212 | Right inferior parietal lobule (IPL), Left anterior cingulate cortex (ACC) |
| 203, 212 | Right medial cingulate cortex (MCC), Left anterior cingulate cortex (ACC) |
| 204, 212 | Right supramarginal gyrus (SupMG), Left anterior cingulate cortex (ACC) |
| 126, 214 | Left fusiform gyrus (FG), Left medial frontal gyrus (MFG) |
| 203, 214 | Right medial cingulate cortex (MCC), Left medial frontal gyrus (MFG) |
| 181, 215 | Right medial frontal gyrus (orbital, MFG), Left anterior cingulate cortex (ACC) |
| 189, 215 | Right medial frontal gyrus (MFG), Left anterior cingulate cortex (ACC) |
| 192, 215 | Right inferior parietal lobule (IPL), Left anterior cingulate cortex (ACC) |
| 196, 215 | Right medial frontal gyrus (MFG), Left anterior cingulate cortex (ACC) |

| | |
|---|---|
| 124, 217 | Left parahippocampal gyrus (PHG), Right anterior cingulate cortex (ACC) |
| 189, 217 | Right medial frontal gyrus (MFG), Right anterior cingulate cortex (ACC) |
| 204, 217 | Right supramarginal gyrus (SupMG), Right anterior cingulate cortex (ACC) |
| 215, 218 | Left anterior cingulate cortex (ACC), Right superior frontal gyrus (SFG) |

*Supplement 1.1. Neuropsychological Testing*

First, participants were asked to complete a series of neuropsychological assessments in their pre-treatment session to determine baseline measures of cognition, quality of life, and pain. Selected domains from the NIH Cognition Battery, PROMIS (Patient-Reported Outcomes Measurement Information System; regarding pain interference, pain intensity, etc.) and Neuro-QoL™ (Quality of Life in Neurological Disorders; regarding fatigue, depression, anxiety, etc.) were obtained from the NIH Toolbox and were administered via iPad. Any additional assessments were administered via paper copy. This set of assessments (Supplemental Information, Table 1) was administered to each subject both pre- and post-ACT.

*Supplement 1.2. Neuropsychological Assessment Analysis*

Assessment data were entered into Excel spreadsheets using Qualtrics software (Qualtrics, 2005) for statistical analysis. Analyses were conducted using SAS® v.9.4. (SAS Institute, 2011) to yield measures such as mean, standard deviation, Student's T (Change), Pr > |t|, Wilcoxon Signed Rank, and Pr >= |S| for a total score pre-ACT, total score post-ACT, and the change score between the two. Positive or negative change scores indicated satisfactory results, depending on the specific assessment in question (CESD scores decreasing meant less depression, and AAQ-II scores increasing meant greater feelings of acceptance, e.g.).

*Supplement 1.3. Neuropsychological Assessment and rsfMRI Data Correlation Analysis*

Additionally, the fMRI data set was merged with the neuropsychological assessment data set so that any correlations between change in neural plasticity (functional connectivity measures) and change in neuropsychological health indicators (behavioral measures) could be analyzed pre- and post-ACT. An Excel spreadsheet was compiled of each subject's pre- and post-ACT values for all significant edge connections (56 total; derived from an NBS analysis between DMN, FPN, and SN with a threshold of t>2.1 instead of t>2.5; see Supplemental Information, Table 5) as well as certain significant change scores (9 reported; AAQ-II, BPI, CESD, FFMQ, PCL-5, Neuro-QoL Depression, Neuro-QoL Fatigue, Neuro-QoL Satisfaction with Social Roles and Activities, and NIH Toolbox Pattern Comparison Processing Speed; the CPAQ was not used). These 9 tests (which previously yielded significant change results; see Supplemental Information, Table 6) were selected for a more cohesive understanding of brain region activation as it corresponds to chronic pain-specific assessments. SAS® v.9.4 was again used to derive correlational data (pairwise Pearson correlation, R, and p value) between functional connectivity changes and neuropsychological assessment changes. For the first pass, all scores with a $p < 0.10$ were selected. The list was then cut down to seven assessment measures and 15 functional connectivity measures. For the second pass, all scores with a $p < 0.0071$ were selected (based on Bonferroni equation for multiple comparison correction), in addition to a $p < 0.05$.

*Supplement 1.4. Neuropsychological Assessment Data Change Results*

Improvements were found based on the assessment data from pre- to post-ACT. Of the administered tests, approximately half were found to have significant change scores (Supplemental Information, Table 6). Negative values for the following assessments represent: lower levels of pain severity (BPI), depression (CESD, Neuro-QoL Depression), PTSD (PCL-5), and fatigue (Neuro-QoL Fatigue). Positive values for the following assessments represent: higher levels of chronic pain acceptance and action (AAQ-II, CPAQ), mindfulness (FFMQ), processing speed (NIH Toolbox PCPS), and satisfaction with social roles (Neuro-QoL Satisfaction with Social Roles & Activities).

**SI Table 6.** Statistically significant change scores of assessments administered.

| **Neuropsychological Assessment (Change Score)** | **S** | **Pr >= \|S\|** | **T** | **Pr > \|t\|** |
|---|---|---|---|---|
| AAQ-II | 20 | 0.0156 | | |
| BPI | -16.5 | 0.0234 | | |
| CESD | -18.5 | 0.0273 | | |
| CPAQ | 20 | 0.0156 | | |
| FFMQ | 20.5 | 0.0117 | | |
| PCL-5 | -21.5 | 0.0078 | | |
| Neuro-QoL Depression | -17.5 | 0.0352 | -2.37633 | 0.0448 |
| Neuro-QoL Fatigue | -17 | 0.0156 | -3.16337 | 0.0133 |
| Neuro-QoL Satisfaction with Social Roles & Activities | 22.5 | 0.0039 | 2.35737 | 0.0461 |
| NIH Toolbox Pattern Comparison Processing Speed (PCPS) Test: | - | - | - | - |
| *Age Corrected Standard Score* | 22.5 | 0.0039 | 7.49688 | <0.0001 |
| *Computed Score* | 22.5 | 0.0039 | 6.11596 | 0.0003 |
| *Fully Corrected Score* | 22.5 | 0.0039 | 8.176373 | <0.0001 |
| *Item Count* | 22.5 | 0.0039 | 5.962922 | 0.0003 |
| *National Percentile (Age Adjusted) Score* | 22.5 | 0.0039 | 4.625803 | 0.0017 |
| *Uncorrected Standard Score* | 22.5 | 0.0039 | 6.189544 | 0.0003 |

**Note.** S: Wilcoxon Signed Rank; T: Student's T (Change). T scores were not reported for assessments administered by paper (AAQ-II, e.g.) because normality could not be assumed. Change scores represent the difference in score between the first and second timepoints (negative S and T values indicate that the score post-ACT was lower than pre-ACT, while positive S and T values indicate post-ACT scores higher than pre-ACT).

*Supplement 1.5. Neuropsychological Assessment and rsfMRI Data Correlation Results*

To further the investigation, the behavioral data and the resting state fMRI data were run together to search for any correlational relationships. Through multiple passes, the initial edges and assessments were narrowed down to 15 edges and seven assessments. The final pass led to the correlations found in Supplemental Information, Table 7.

13 edges were included in significant ($p < 0.05$) correlations with six assessments (left column and top row in Supplemental Information, Table 2). The first assessment (AAQ) was correlated with one edge representing the functional connection between right AG and left ACC. The second assessment (BPI) was correlated with five edges representing the functional connections between left PHG and right IPL, right SFG and right PCG, right SFG and right IFG, right MFG and left ACC, and right SupMG and right ACC, respectively. The third assessment (FFMQ) was correlated with two edges representing the functional connections between left PHG and left PCG, and left PHG and right SFG, respectively. The fourth assessment (NQ DEP) was correlated with two edges representing the functional connections between right MFG and left ACC, and right PCG and right Ins, respectively. The fifth assessment (NQ SSR) was correlated with three edges representing the functional connections between right MTG and left PHG, left PHG and right SFG, and right IFG and right MFG, respectively. The sixth assessment (NIH Toolbox PCPS) was correlated with one edge representing the functional connection between left MCC and left ACC.

So, the significant changes in functional connectivity, namely within cingulate cortex and parahippocampal, precentral, and frontal gyri, were correlated with the significant changes in assessments regarding pain, social satisfaction, depression, and processing speed pre- to post-ACT. To correct for multiple comparisons, the data were assessed using a p value of 0.0071 (calculated by dividing 0.05 by the final number of assessments included, seven). Only one correlation involving the right angular gyrus and left anterior cingulate was significant using the Bonferroni method described (starred in Supplemental Information, Table 7).

**SI Table 7.** Correlations between significant assessment and edge scores.

| | AAQ | BPI | FFMQ | NQ_DEP | NQ_SSR | PCPS |
|---|---|---|---|---|---|---|
| L MCC, L ACC | | | | | | 0.77662<br>**0.0138** |
| R AG, L ACC | 0.86811<br>**0.0024*** | | | | | |
| R MTG, L PHG | | | | | -0.74181<br>**0.0221** | |
| L PHG, L PCG | | | -0.81066<br>**0.008** | | | |
| L PHG, R SFG | | | -0.68596<br>**0.0413** | | -0.81151<br>**0.0079** | |
| L PHG, R IPL | | -0.77014<br>**0.0152** | | | | |
| R IFG, R MFG | | | | | -0.6976<br>**0.0367** | |
| R SFG, R PCG | | -0.7447<br>**0.0213** | | | | |
| R SFG, R IFG | | -0.69481<br>**0.0378** | | | | |
| R MFG, L ACC | | | | -0.66811<br>**0.0492** | | |
| R MFG, L ACC | | -0.76687<br>**0.0159** | | | | |
| R SupMG, R ACC | | -0.79017<br>**0.0113** | | | | |
| R PCG, R Ins | | | | 0.67389<br>**0.0466** | | |

**Note.** All significant correlations between edge (blue) and assessment (orange) change scores. P values are bold, listed below the R values. *This represents the only value (0.0024) that is < 0.0071 of the significant correlations.

*Supplement 1.6. Main Findings in Neuropsychological Assessment Data*

Behavioral data were collected using select domains for the NIH Cognition Battery, PROMIS and Neuro-QoL on the NIH Toolbox as well as physical paper surveys (Supplemental Information, Table 1). Approximately half of all of the assessments changed significantly. All of the significant change scores demonstrated improved scores for the nine participants (Supplemental Information, Table 6). All participants exhibited lower levels of pain severity, depression, PTSD, fatigue, as well as higher levels of chronic pain acceptance, mindfulness, processing speed, and satisfaction with their social roles. This indicates that the ACT enhanced quality of life in very important aspects of the overall perception of chronic pain condition. Chronic pain sufferers form negative life views more often than not. However, this study and previous studies point to ACT as a meaningful, non-invasive, life-enhancing treatment for various chronic pain conditions.

Previous studies of the behavioral outcomes post-ACT have found similar improvements for daily physical and social functioning in addition to alleviated psychological flexibility by improved depression and mindfulness scores (Scott et al., 2016; Hann & McCracken, 2014; Veehof et al., 2011; McCracken & Gutiérrez-Martínez, 2011; Wicksell et al., 2013; Vowles et al., 2011; Wicksell et al., 2010; Vowles et al., 2014). Others have demonstrated a resultant improvement of mental health measures of acceptance and value-based action, which would support the change score results on the AAQ-II in the current study (Fledderus et al., 2010). Dahl and Lundgren have demonstrated this acceptance to be linked to lower self-rated depression scores, greater physical and social abilities, and less pain avoidance, which would further support the change scores on the CESD, Neuro-QoL Depression, and Satisfaction with Social Roles assessments in the current study (Dahl & Lundgren, 2006).

*Supplement 1.7. Main Findings in Correlation Analysis Data*

Because of the significant findings of the behavioral assessment analysis and the rsfMRI graph theory analysis, it was critical to then investigate if the two independent findings were correlated to each other. This additional correlational analysis was conducted (ultimately) using seven behavioral assessment measures, 15 rsfMRI functional connectivity measures (edges), and the SAS® v.9.4 software. Pairwise Pearson correlations exhibited 14 significant correlations that were either strong (|0.6| – |0.8|) or very strong (> |0.8|) correlations (Table 2). Within the 14 correlations, four of the assessments (BPI, FFMQ, NQ DEP, and NQ SSR) correlated with more than one edge. This indicates important functional connections related to the improved assessment scores. Specifically, the BPI scores were significantly correlated with five connections (involving frontal gyri, cingulate, and more), demonstrating multiple important interactions between ACT-induced neural plasticity and subjective perception of pain severity.

Of importance is the connection between the right angular gyrus and the left anterior cingulate that was significantly correlated with the AAQ-II when correcting for multiple comparisons (using the Bonferroni method). At a p value of 0.0024 and R = 0.86811, this is a very strong correlation. The anterior cingulate has been previously linked to the cognitive and emotional regulation of pain processing as well as response selection, as opposed to aspects of pain intensity (Feliu Soler et al., 2018; Peyron et al., 2000). In future investigations, the anterior cingulate and angular gyrus should be considered as potential critical underlying neural mechanisms. In addition, the behavioral measure for acceptance and action surrounding chronic pain should be studied more in depth.

Previous studies have also shown similar neurological interactions and revealed the importance of their connections. In numerous other ACT investigations with a chronic pain population, the medial frontal gyrus (found to have numerous significant correlations in the current study) has shown similar deactivation patterns (Smallwood et al., 2016, Feliu Soler et al., 2018; Taskiran Sag et al., 2017). The medial frontal gyrus is namely responsible for assessing the risk of chronic pain as it manifests, possibly even before the chronicity is established (Mano et al., 2018). When connections between the prefrontal cortex and anterior cingulate have been shown, those connections have been attributed to attentional and memory network activation as a response to the painful stimuli (Peyron et al., 2000). In discussing the anterior cingulate, it is important to note the region's probable link to anxiety and depression (as depression measures were decreased in the current study; Smallwood et al., 2014). Additional studies have also demonstrated activation changes in the anterior cingulate after ACT interventions, further indicating its level of interaction (Feliu Soler et al., 2018; Taskiran Sag et al., 2017).

### Supplemental Information References

Dahl, J., & Lundgren, T. (2006). *Acceptance and commitment therapy in the treatment of chronic pain*. 36. http://dx.doi.org/10.1016/B978-012088519-0/50014-9

Feliu Soler, A., Montesinos, F., Gutiérrez-Martínez, O., Scott, W., McCracken, L., & Luciano, J. (2018). Current status of acceptance and commitment therapy for chronic pain: A narrative review. *Journal of Pain Research*, *11*, 2145–2159. https://doi.org/10.2147/JPR.S144631

Fledderus, M., Bohlmeijer, E.T., Smit, F., & Westerhof, G.J. (2010). Mental health promotion as a new goal in public mental health care: A randomized controlled trial of an intervention enhancing psychological flexibility. *American Journal of Public Health*, *100*, 2372–2372. https://doi.org/10.2105/AJPH.2010.196196

Hann, K.E.J., & McCracken, L.M. (2014). A systematic review of randomized controlled trials of Acceptance and Commitment Therapy for adults with chronic pain: Outcome domains, design quality, and efficacy. *Journal of Contextual Behavioral Science, 3*, 217-227. https://doi.org/10.1016/j.jcbs.2014.10.001

Mano, H., Kotecha, G., Leibnitz, K., Matsubara, T., Nakae, A., Shenker, N.,…Seymour, B. (2018). Classification and characterisation of brain network changes in chronic back pain: A multicenter study. *Wellcome Open Research, 3*. https://doi.org/10.12688/wellcomeopenres.14069.2

McCracken, L.M., & Gutiérrez-Martínez, O. (2011). Processes of change in psychological flexibility in an interdisciplinary group-based treatment for chronic pain based on acceptance and commitment therapy. *Behaviour Research and Therapy, 49*, 267–274. https://doi.org/10.1016/j.brat.2011.02.004

Peyron, R., Laurent, B., & García-Larrea, L. (2000). Functional imaging of brain responses to pain. A review and meta-analysis (2000). *Neurophysiologie Clinique = Clinical Neurophysiology, 30*, 263–288. https://doi.org/10.1016/s0987-7053(00)00227-6

Power, J.D., Cohen, A.L., Nelson, S.M., Wig, G.S., Barnes, K.A., Church, J.A.,…Petersen, S.E. (2011). Functional network organization of the human brain. *Neuron*, *72*, 665–678. https://doi.org/10.1016/j.neuron.2011.09.006

Scott, W., Hann, K.E.J., & McCracken, L.M. (2016). A comprehensive examination of changes in psychological flexibility following acceptance and commitment therapy for chronic pain. *Journal of Contemporary Psychotherapy*, *46*, 139-148. https://doi.org/10.1007/s10879-016-9328-5

Smallwood, R.F., Hutson, R.M., & Robin, D.A. (2014). Neuroimaging connectivity analyses and their application in psychiatric research. *Pathobiology of Human Disease*, 2522–2537. https://doi.org/10.1016/B978-0-12-386456-7.05217-5

Taskiran Sag, A., Ceylan Has, A., Oztekin, N., Temucin, C.M., & Karli Oguz, K. (2017). Tracking pain in resting state networks in patients with hereditary and diabetic neuropathy. *Noro Psikiyatri Arsivi, 56*, 92-98. https://doi.org/10.5152/npa.2017.22660

Veehof, M.M., Oskam, M-J., Schreurs, K.M.G., & Bohlmeijer, E.T. (2011). Acceptance-based interventions for the treatment of chronic pain: A systematic review and meta-analysis. *Pain*, *152*, 533–542. https://doi.org/10.1016/j.pain.2010.11.002

Vowles, K.E., McCracken, L.M., & O'Brien, J.Z. (2011). Acceptance and values-based action in chronic pain: A three-year follow-up analysis of treatment effectiveness and process. *Behaviour Research and Therapy, 49*, 748-755. https://doi.org/10.1016/j.brat.2011.08.002

Vowles, K.E., Witkiewitz, K., Sowden, G., & Ashworth, J. (2014). Acceptance and commitment therapy for chronic pain: Evidence of mediation and clinically significant change following an abbreviated interdisciplinary program of rehabilitation. *The Journal of Pain, 15*, 101–113. https://doi.org/10.1016/j.jpain.2013.10.002

Waller, N.C., Ray, K.L., Meier, S.K., Aytur, S.A., & Robin, D.A. (Unpublished results). Regional brain activation in chronic pain: A functional connectivity meta-analysis with healthy controls and chronic pain patients.

Wicksell, R.K., Kemani, M., Jensen, K., Kosek, E., Kadetoff, D., Sorjonen, K.,…Olsson, G.L. (2013). Acceptance and commitment therapy for fibromyalgia: A randomized controlled trial. *European Journal of Pain, 17*, 599–611. https://doi.org/10.1002/j.1532-2149.2012.00224.x

Wicksell, R.K., Olsson, G.L., & Hayes, S.C. (2010). Psychological flexibility as a mediator of improvement in acceptance and commitment therapy for patients with chronic pain following whiplash. *European Journal of Pain, 14*, 1059. e1011–1059.e1051. https://doi.org/10.1016/j.ejpain.2010.05.001
